# How are declarations of interest working? A cross sectional study in declarations of interest in medical practice in Scotland and England in 2020/2021

**DOI:** 10.1101/2022.03.11.22271902

**Authors:** M McCartney, RB Hartman, H Feldman, R MacDonald, F Sullivan, C Heneghan, C McCutcheon

## Abstract

**Objective:** To understand arrangements for doctors’ declarations of interest in Scotland and England in the context of current recommendations.

**Design:** Cross sectional study of a random selection of NHS hospital registers of interest by two independent observers in England, all NHS Boards in Scotland, and a random selection of Clinical Commissioning Groups (CCGs) in England.

**Setting:** NHS Trusts in England (NHSE), NHS Boards in Scotland, CCGs in England, and private healthcare organisations.

**Participants:** Registers of declarations of interest published in a random sample of 67 of 217 NHS Trusts, a random sample of 15 CCGs of in England, registers held by all 14 NHS Scotland boards, a purposeful selection of private hospitals/clinics in the UK.

**Main Outcome Measures:** Adherence to NHSE guidelines on declarations of interests, and comparison in Scotland.

**Results:** 76% of registers published by Trusts did not routinely include all declaration of interest categories recommended by NHS England. In NHS Scotland only 14% of Boards published staff registers of interest. Of these employee registers (most obtained under Freedom of Information), 27% contained substantial retractions. In England, 96% of Clinical Commissioning Groups published a Gifts and Hospitality register, with 67% of CCG staff declaration templates and 53% of governor registers containing full standard NHS England declaration categories. Single organisations often held multiple registers lacking enough information to interpret them. Only 35% of NHS Trust registers were organised to enable searching. None of the private sector organisations studied published a comparable declarations of interest register.

**Conclusion:** Despite efforts, the current system of declarations frequently lacks ability to meaningfully obtain complete health care professionals’ declaration of interests.

## Introduction

Methods of recording, disclosing and managing conflicts of interest in healthcare have been vigorously debated in recent decades. These have cumulated in a variety of ‘Sunshine Acts’ enacted between 2012-2018 in North America, Australia and some European countries (1), but notably not the UK. These Acts variously mandate public disclosure of healthcare professionals’ affiliations with industry, either by the individuals, or by pharmaceutical or other companies.

Declarations of interest are also now commonly required by employers, journals, conferences, guideline or other committees, but this was not always the case. Until the 1980s, declarations made by journal authors were haphazard and voluntary (2). After a series of medical frauds and misconduct in the USA, the House Science and Technology Committee published “Is Science for Sale? Conflicts of Interest vs. the Public Interest” (3). The Committee heard how financial links between companies, research institutes and universities were a ‘recipe for conflicts of interest’ and how ‘very few scientists would admit that the commercial associations have affected them personally’. Subsequently, medical organisations became more interested in requesting conflicts of interest from their contributors. In 1993, The International Committee of Medical Journal Editors wrote that authors must “recognise and disclose financial and other conflicts of interest that might have biased their work. They should acknowledge in the manuscript all financial support for the work and other financial and personal connections to the work” (4). This was later formalised with explicit instructions for disclosure (5).

Despite the ubiquity of requests to disclose declarations, there are numerous unresolved tensions, including questions of composition, purpose, and utility. This is reflected in the names given to disclosures required in situations ‘where the impartiality of research may be compromised because the researcher stands to profit in some way from the conclusions they draw” (6). For example, a disclosure of “conflicts of interest” requires insight and judgement as to when disclosure is applicable, as does a requirement to state “dual commitments” “competing interests” or “conflicting loyalties”. A request for a “declaration of interests” does not imply that disclosures are conflicts; merely that they exist: some organisations ask for “potential” rather than actual, or perceived actual, conflicts. Critical are findings that while doctors recognise that other doctors, subject to gifts from industry, may be conflicted in their judgements, they tend to feel they themselves are immune to the same influences (7). This suggests questionable insight, and infers that judgements over when an interest constitutes a conflict are best made by an independent observer. This would therefore support disclosures of “interests” rather than “conflicts”, which reflects NHS England practice.

Some have argued that disclosure is a fallacy, as disclosure cannot negate a conflict (8). However, the inference that information about conflicts may as well be left undisclosed opposes many professional regulators’ expectation of transparency (9). Indeed, a lack of candour and openness in management of conflicts is described as a risk to professional trust and credibility by some medical organisations (10). This is concerning due to the evidence around the conflicts of interest in patient care. For example, financial interests are associated with potential bias in systematic reviews (11) and distorted outcomes from drug and device studies (12). Pharmaceutically sponsored medical education is associated with more expensive, poorer quality prescribing (13), and guidelines for use of opioids in non cancer pain are prone to bias due to authors’ conflicts of interests, now implicated in the opioid crisis (14). Non disclosure would make such associations more difficult to find: this knowledge does potentially allow the journal reader, to some extent, make judgements on their relevance and potential impact (15). Some organisations have gone further, for example, Open Payments (16), and Dollars for Docs (17) have encouraged citizens to consider whether their healthcare provider is be conflicted and to search individual practitioner records. It is unknown whether this assists patients to make better quality decisions, and whether it increases or decreases trust in the medical profession - whether justified or not. However, without disclosure, it is not possible to appropriately remove or recuse conflicted individuals from inappropriate positions of power.

What should be disclosed, and where, is a matter of debate. The ICMJE set a rare exception of an internationally agreed standard. In the UK, the dominant medical employer is the tax funded National Health Service. The UK Government in both 2005 and 2021 rejected proposals for central, standardised registers of staff declared interest in favour of holding them at local level (18). However, there was demonstrable poor practice in declarations published by hospital employers in 2015-16 (19). In 2016, a review in NHS England recommended improvements in declarations for hospital employers and Clinical Commissioning Groups (CCGs, who direct funding in primary care) in England, issuing templates for defined staff groups to record interests (relevant shareholding and ownership interests, professional/personal financial/non financial direct or indirect interests; actions to be taken in respect of a conflict) and instructions for completion of gifts and hospitality registers. These were intended to ensure consistent declarations, to be published “in a prominent place” online, and be “accessible and contain meaningful information”. Information could be exceptionally redacted when “real risk of harm or is prohibited by law” (20). CCGs received similar, statutory guidance regarding declarations in 2017. Scotland has had no similar recent review. In requesting NHS Scotland current policies for staff to declare interests, we were asked to apply via a Freedom of Information request and advised that NHS Circular No 1989 (21), which applies to medical and non-medical staff, was still extant. It states that staff must notify their employer if they have a relationship with a current or likely future contractor, and “establish and maintain registers both of the financial interests of any staff involved in purchasing/commercial policy and of any ‘gifts or considerations’ received by staff from any commercial sources.” A further circular in 1994 offers guidance for declarations for Board Members (22). While NHS England recommends annual publication of relevant registers on a prominent place on their website, there is no equivalent national statement for NHS employees in Scotland. Declarations to employers in Scotland is therefore inferred to involve self judgement to declare conflicts, not interests. However, individual Board policy may state that staff interests should be declared and/or be open to public access.

There is therefore a working assumption that disclosures of interest are necessary and should contain enough information for others, whether professional or lay, to interpret their meaning. However, we do not know whether guidance to NHS England employers has resulted in registers of declarations compliant with guidance, what current declarations practice is within NHS Scotland, how healthcare professionals have responded to calls for their declarations, and whether this has resulted in interpretable information for lay or professional readers.

## Study Objective

In the United Kingdom, The Independent Medicines and Medical Devices Safety Review (2020), like the Health Select Committee before it (2005), recommended doctors make a statutory declaration of their interests on a central register (23,24). This proposal has been rejected by government who instead continue to recommend locally made declarations. However it is uncertain whether these are fit for purpose in describing actual or potential conflicts of interest, and enable others to make an informed judgement on their relevance and potential effects. We therefore examined current practice of public declaration of interests in hospitals in England and Scotland, and CCGs in England, along with a purposeful selection of private clinics and hospitals. Due to resource limitations we excluded Wales and Northern Ireland from our study. We judged declarations in NHS England, Scotland and the private sector against contemporary NHS guidance in England. We aimed to understand current practice in declaring interests in the UK by assessing the compliance of employers in publishing registers of interest as recommended by NHS England guidance, how employees have responded to these requests, and the organisational effectiveness of declarations of being available and containing enough information to interpret them.

## Methods

We examined the current practice of public declaration of interests in hospitals in England and Scotland, CCGs in England, and a purposeful selection of private organisations. Searches, as described below, were performed between April and July 2021. Data extraction was performed by two researchers independently and disagreements resolved by discussion.

This research is part of a wider suite of work into declarations of interest, which has a Patients/ Citizens panel (25). This project was suggested and discussed by and with members, who contributed in particular to what would contribute to a meaningful declaration of interest; and what private clinics and hospitals we should review.

### NHS Trusts and Boards

There were 217 NHS Trusts in England as of April 2021. We sampled 67 to give 95% confidence, with 10% margin of error, in in locating representative registers. (Initial randomisation included the Mid Staffs and Nightingale Trusts: the former had been included on the official NHS Trust list erroneously, as it was dissolved at the time of research: the latter was temporary, using seconded staff to deal with Covid-19 pandemic pressures. We therefore did not think it fair to judge by usual standards; these Trusts were removed and replaced.) In Scotland, there are 14 Health Boards.

Because this is the first investigation of this type we are aware of, and the smaller number, we included all.

We searched organisations” websites for registers of

- Board, governor, member or decision maker
- Gifts and Hospitality
- Staff

If each of these registers were not immediately apparent, or combined, Trust sites were searched systematically (appendix 1).

Sampling of NHS England Trust websites used a random number generator, set against an alphabetical list of Trusts. In each case, the “test” was of the recording process used to generate declarations, not the content of individuals’ responses. We gathered specific data for each organisation (appendix 2) using a search strategy (appendix 3) assessing whether the template adhered to all requirements set by NHSE (appendix 4). These were assessed by two researchers independently and disagreements resolved by discussion.

Contacting Trusts: If neither researcher located each register, we emailed the media department of the Trust (appendix 5) requesting any publicly available register of interests. If there was no reply in 7 days we wrote again and recorded outcomes.

### NHS Boards - Scotland

Preliminary searches found few public registers. Because we were not aware of any previous systematic analysis of registers held by NHS Scotland boards, we used different methods in order to understand current practice.

In assessing current practice in Scotland we were interested in whether staff declarations were being requested and if so, published. We used the same searching methodology as used in English Trusts. In the absence of national guidance to publish declarations, we sent FOI requests to each Board (appendix 6) requested published and unpublished registers, and recorded all. We assessed register categories against NHSE guidance in order to compare whether Scottish declarations captured more, less, or the same data as in England.

A 20% sample of these were checked with a second researcher.

### CCGs - England

In 2020 there were 191 Clinical Commissioning Groups (CCGs) in England, merging into 135 CCGs in 2020-21, many containing multiple entities with some shared and some separate registers. We altered our search strategy in accordance with NHS England guidance on register templates (Appendix 7,8). More registers than anticipated were found (for example, 7 registers in a single CCG). Because of resource limitations and an unknown denominator, we stopped searching after analysis of 68 registers in 15 CCGs repeatedly obtained similar results. All CCG entities had to contain relevant registers to meet the criteria.

### Detailed Analysis of a sample of NHS England registers

NHS England guidance states declarations should “contain enough information to be meaningful (e.g. detailing the supplier of any gifts, hospitality, sponsorship, etc). That is, the information provided should enable a reasonable person with no prior knowledge should be able to read this and understand the nature of the interest.” We discussed this with the patient group. We assessed financial declarations of decision makers using the framework of understanding ‘who/what/where/ when’ of financial transactions, given the evidence of impact, and ability to more consistently demarcate findings. Individual declarations were randomly selected from Trusts, themselves randomly selected. We intended to search a larger sample, but given the lack of clear denominator (staff members who made a financial DOI), variability in reporting systems and disclosures made, and limited resources, we stopped assessing entries after all found regular deficiencies. If a declaration had no financial component, N+1 was used. Data was extracted from Trusts using ‘mydeclarations’ (a proprietary software brand used by many Trusts (26) to locate individuals. Registers with fewer than 3 entries were disregarded.

We assessed :

1) Type of financial interest

2) Whether “a reasonable person with no prior knowledge should be able to read this and understand the nature of the interest”: who paid the money, what was the quantity, why and when was it paid, and what action was intended to mitigate it.

Pharmaceutical company declarations were cross checked on the voluntary disclosure website Disclosure UK. This was a search of staff COI registers. If the declaration was a ‘gift’ or ‘hospitality’, and the Trust had a Gifts and Hospitality register, this was searched to assess consistency.

### Private sector

With the patient group, we considered how to identify which organisations to examine. We assessed declarations on the five largest private hospital chains in the UK and the five top Google searches for ‘private health clinics uk’, including promoted sites, if not already captured. The Private Healthcare Market Investigation Order 2014 (27) states that hospital operators must publish details of clinicians with shares or financial interests in the hospital or equipment. We assessed websites for statements relating to any disclosure of interests.

## Results

### England - NHS Trusts

We found a total of 162 registers published by 67 Trusts (median 2, range 1-4, Appendix 9). Some registers were absent: We could not find all the relevant registers on 25 Trust websites: direct contact produced two further registers.

Of these, 21% of board registers and 65% of governor registers were absent (Table 1); 43% of Trusts published a decision maker register and 23% an ‘all staff’ register. 46% of Trusts published a separate gifts and hospitality register, and of those that did not, 28% published it in a combined register elsewhere. Of 162 registers analysed (Table 2), 29% were at least 14 months old, 24% contained all the categories recommended by NHS England, and 35% were searchable either by alphabet or online search box. 69% of registers held by Trusts enabled a declaration of no interests to be made, and 26% asked for quantification of financial interests.

### Scotland - Board Registers

All NHS Scotland Board members published registers of interest, however only one published a staff Declaration of Interest register. Seven of 14 Boards published a Gifts and Hospitality register (Table 3). Two registers obtained under FOI (appendix 10) contained all NHS England template categories on Gifts and Hospitality. Of 33 registers available to inspect, 9 contained substantial retractions, e.g. names of staff or types of interest for hospitality or sponsorship. Numerous examples of hospitality funded for by multiple pharmaceutical companies for ‘Grand Rounds’ or ‘junior doctors meeting’ had no further detail supplied. Other examples of difficult to interpret information were declarations such as ‘honorarium’, or ‘consultancy’ with no further detail supplied. Two health boards did not provide staff Register of Interests under FOI, with one Board saying they were recorded at “local level”, not centrally recorded and spread across hundreds of departments; the other was described as not being in an easily retrievable format. Two registers were empty but dated, hence the different denominators in some analysis. All had board member registers, and registers for Gifts and Hospitality, sometimes included in a Staff Register. One Board held a register purely for ‘sponsorship’. Eight boards had a register for all staff declarations of interest, and one other for senior decision makers/managers only.

### Detailed analysis of NHS England registers

On detailed examination (appendix 11), we found instances of Hospitality declared on a register of interest, but not on the gifts and hospitality register (Table 4). Declarations made on a proprietary website (mydeclarations.co.uk) included template questions including date, sponsor, description, value, and recipient. However, this still allowed for error. For example, a nurse consultant declared income for paid talks as ‘sponsorship’ when this should have been categorised as outside (freelance) employment. We found instances where declarations had been made on DeclarationsUK (28) an industry open declarations initiative, but not on employers websites. The proprietary website (mydeclarations) contained multiple categories, but not always a statement of actions taken in order to mitigate potential conflicts.

## Private sector

No public gifts and hospitality register was located on any assessed website (Table 5). Each published necessary Competition and Markets Authority statements relating to potential conflicts with the hospital (e.g. shares and equipment ownership), but not conflicts of interest more broadly, either on each consultants’ profile, or a central register. Each hospital chain/private clinic was contacted. Three replied confirming they published no other register. There was no reply after two contacts with the other hospitals. Of the 5 private health clinics listed first in a Google search for ‘private health clinic’, three provided a CMA statement; none provided Gifts, Hospitality, or Conflicts of Interest registers (Table 6, appendix 12).

### Clinical Commissioning Groups

There were very high levels of Gifts and Hospitality register publication (Table 7) and over two thirds used NHS England standard template categories. Many governors declared their own GP practice as a potential COI.

## Discussion

There are significant problems with current practice which result in frequently ineffective declaration of interests, not simply in terms of composition, but also in organisation and utility. Healthcare organisations commonly hold multiple registers of declarations, which can include Board members, Governors, decision making staff, all staff, gifts and hospitality, sponsorship, and for Competition and Markets Authority stipulations. These may be combined or contained within separate documents. Some were absent, and while many organisations filed consistent, visible declarations, some were contained in minutes of meetings, individual biographies, or only on direct request of Freedom of Information request.

Even if organisations were complaint with publishing standard templates, meanings were often opaque. For example, lunches were frequently declared as paid by numerous companies for ‘grand rounds’, without further detail. Registers could state ‘pharmaceutical shares’ ’honorarium’ or ‘director of a management consultancy’, but without information to interpret the nature and extent of conflicts. Only 10% of registers published by Trusts contained specific action to be taken regarding each type of declaration. Declarations were noted containing a self-justification, for example “I have completed the gift register for all of the talks and other company links I have. These are all for individual episodes and I do NOT have any long term / contract based contracts with any pharma”. Based on discussions with our patient group, we provide examples of what we judged as high quality statements (the who/what/when/why of the potential conflict), and low quality statements (unlikely to have information to reasonably interpret the disclosure) (Box 1 and 2).

Roughly one third (35%) of registers from Trusts were searchable, mainly alphabetically. Some Trusts used specifically designed, searchable software allowing continuous rolling declarations.This had the potential to include all categories of professionals and types of register but was rarely used as such, with some categories unused, necessitating yet further registers. Additionally, wording on registers may cause confusion over whether a declaration was absent due to nothing to declare, or due to failure to comply with completion. Of registers published by Trusts, 69% made it possible for staff to make a positive declaration of no interests. Private clinics/ hospitals had similar issues: the Competitions and Marketing Authority statement was absent on 2 websites examined, but statements of interest included only those in relation to CMA stipulations and not more broadly, consequently potentially appearing complete when it is not.

There were multiple instances of disclosure of non-recurrent small gifts from patients, e.g. knitted items, hand made jewellery, or flowers, with professionals justifying acceptance lest they be seen as rude. It seems inefficient and wasteful to scrutinise this when known concerns - particularly financial interactions with industry - are difficult to find and interpret, or not published. Further, governors of CCGs declaring where they are registered as patients is a potential breach of their privacy and security.

## Conclusion

Despite the creation of thousands of registers of interest in the UK, locating and interpreting them remains problematic with an organisationally chaotic system limiting their utility and efficiency. Most Trusts are not following NHS England guidance on publishing declarations. It may be better to concentrate greater efforts into making thorough, interpretable, declarations at high risk of producing conflicts, including sponsored educational events, while eliminating the need to stop ‘over-declaring’ low value, non recurrent gifts. It is important to consider what the purpose of declarations are: if others are expected to draw judgements on their potential impact, appropriate information has to be contained to achieve this. It is unknown whether registers are useful to the public or professionals.The lack of oversight and accountability of the current system is concerning.

## Supporting information

Box 1 and 2

## Data Availability

All data available have been uploaded.

## Ethics

We discussed this project with the University of St Andrews medical school ethics committee and they confirmed that no ethical permissions were required.

