## Appendix 1-8 for "How are declarations of interest working? A cross sectional study in declarations of interest in medical practice in Scotland and England in 2020/2021"

**Appendix 1- Search strategy for NHS Trust registers**

1. Use interface to navigate to

a. About us > FOI > Publication Scheme > Lists and registers
b. About us > Board members > declarations
c. About us > Council of Governors > declarations
d. About us > Statutory information
e. About us > Compliance statements

2. Website (5 pages) and Google search (first page) for

f. Declaration
g. Conflict
h. Interest
i. Register
j. Hospitality
k. Gifts

| **Appendix 2- Analysis of Registers of NHS Trusts** | |
| --- | --- |
| 1) Title of register(s): which groups are included? | Record each register by grouping e.g. board members/Governors/all staff/Gifts and Hospitality |
| 2) Use of standardised official NHS England guidance template for Trust or CCG? | Format as per <https://www.england.nhs.uk/ourwork/coi/> template |
| 3) Ability to search either alphabetically or in search box? | Yes/No |
| 4) A “nothing to declare” column, or an example of a nil/no declaration to make? | Yes/No |
| 5) Explicit categorisation of types of interest, e.g. financial, loyalty by column? | Yes/No |
| 6) Is there a value column? | Yes/No |
| 7) Requested explanation of declared income - e.g what produced or could produce a conflict? Anything more than a description box and a comments box is likely to be sufficient, excluding things covered by other questions | Yes/No - examples of detail/ lack of detail  Who it was from, who went to, what it was used for, and how much. |
| 8) Explicit categorisation of action to be taken as a result of declaration? | Yes/No |
| 9) Gifts and hospitality included in staff register or separately held/not existing? | Yes (on single register) No (separate register or not found ) |
| 10) Incidental findings:   Examples of incomplete, ambiguous or patients identifiable information, or good examples/bad examples of registers, noted redactions/refusals | As noted |

**Appendix 3 - Notes on search strategy for NHS Trusts**

1. We took the title of the register to indicate which staff members were included (Board, decision makers, professional groups).
2. NHS England has published a standard template for Hospital Trusts and
   CCGs  - [https://www.england.nhs.uk/publication/conflicts-of-interest-management-templates](https://www.england.nhs.uk/publication/conflicts-of-interest-management-templates/). NHSE Trusts - https://www.england.nhs.uk/ourwork/coi/[/](https://www.england.nhs.uk/publication/conflicts-of-interest-management-templates/) An affirmative infers inclusion of all columns in the standardised template, i.e. Name/Role/Description of Interest/Dates/Comments. All of these had to be included for the register to be labelled as compliant. Some templates used contained all the requirements of NHSE templates plus additional information.
3. We included any register which allowed searching via alphabetasion or via a search function. Some registers alphabetised using first names only, but these were not deemed ‘searchable’.
4. A negative declaration is any declaration which confirms that no conflict is present, or the ability of a template to declare that there are no interests to declare. This examines the method used by the Trust to ask and record definitive statements of declarations by registrants. If we found a single example of a negative declaration, it was deemed to meet this standard.
5. This is in NHS guidance but not explicitly categorised in NHSE forms as a column.
6. Inclusion of a column with request to disclose either pay bands or amounts were included.
7. NHSE guidance states that professionals should “Provide a description of the interest that is being declared.  This should contain enough information to be meaningful (e.g. detailing the supplier of any gifts, hospitality, sponsorship, etc).  That is, the information provided should enable a reasonable person with no prior knowledge should be able to read this and understand the nature of the interest.”  This should therefore be contained in use of the NHSE template. We also scored Trusts positively if they had a column to systematically document explanations. We have examined this aspect further.
8. Guidance notes say this may be contained in the  ‘comments’ on NHS Trust form, i.e. “This field should detail any action taken to manage an actual or potential conflict of interest.  It might also detail any approvals or permissions to adopt certain course of action”.
9. Some Trusts have amalgamated the gifts and hospitality register and others have not: we recorded whether or not this was the case.
10. Any listing of a professional declining to disclose e.g. ‘permission not given for publication’, any entry where a sentence relating to disclosure was incomplete or information clearly missing. It was not possible to search each register line by line for retractions, due to resources, and some may have been missed.

**Appendix 4 - NHS England Template and Notes**

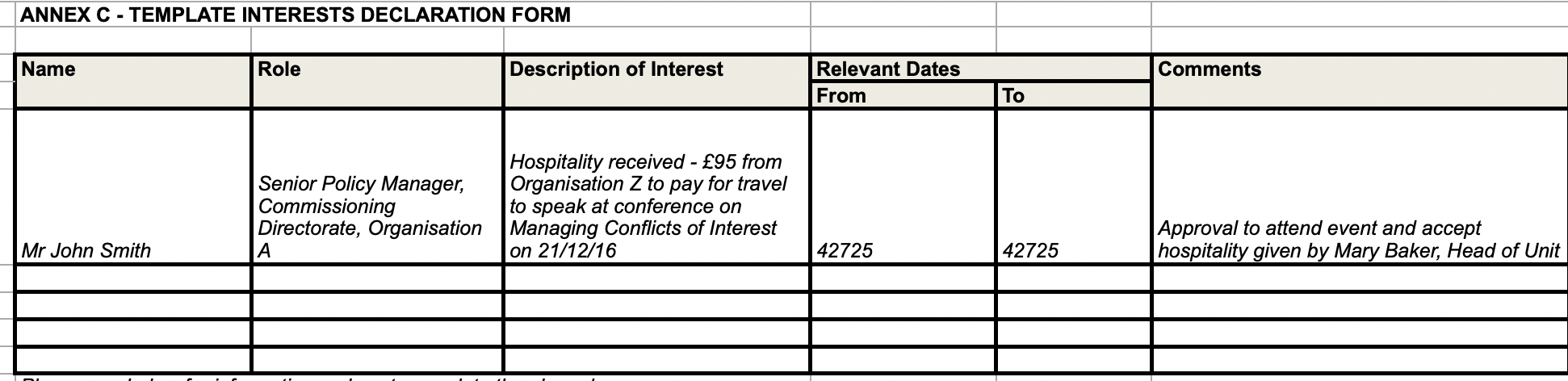

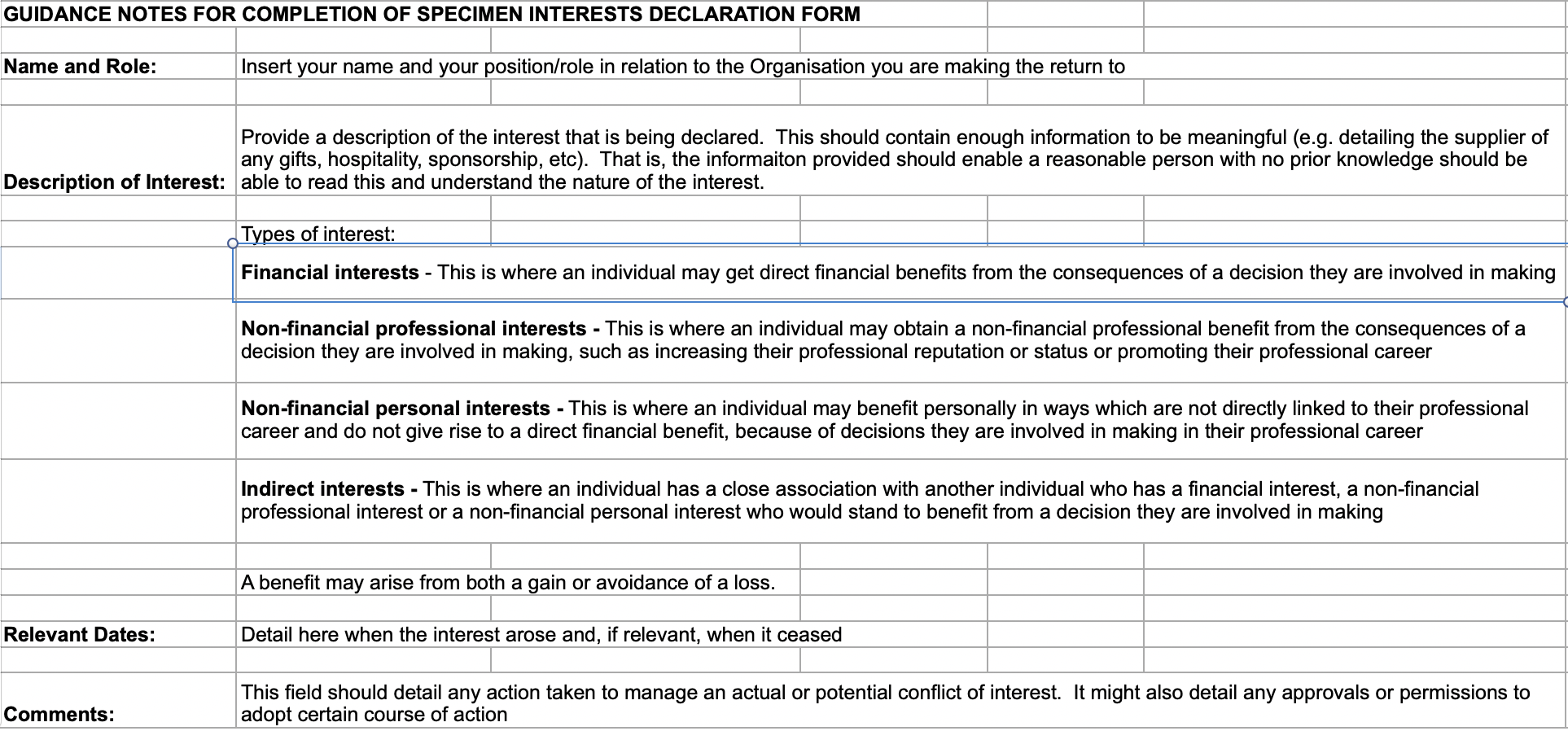

**Appendix 5 - Template letter NHS Trusts**

Dear Communications Team,

We are currently engaged in a research project examining the current practice for declaring interests in NHS Trusts.

I note there is a publicly available register of interests for Board Members/Governors/Other/Gifts and Hospitality register. Is there a publicly available register of interests for senior or all NHS Trust employees? If so, I would be grateful if you could direct me to the URL.

We are only interested in registers already published on your website, not in registers available to the public on request. We appreciate you are under great pressure and your early attention to this would be appreciated. Please get in touch if you require any more information.

Many thanks

**Appendix 6 - Freedom of Information Request to NHS Boards (Scotland)**

Dear Madam/Sir

Request under the Freedom of Information Act

I refer to the  [GEN1989_32.pdf (scot.nhs.uk)](https://www.sehd.scot.nhs.uk/mels/GEN1989_32.pdf) Acceptance of Financial Assistance, Gifts, Hospitality, and Declaration of Interest policy.

This states (paragraph 16) that NHS Boards "The Department expects the Standing Financial Instructions of Health Boards to provide for the declaration of interests by members of staff, who must notify their employer if they have any financial interest in or relationship with a manufacturer, supplier or contractor with whom the Board is entering or is likely to enter into a contractual relationship, and of any financial or other interest which may affect the Board's planning or policy decisions. This includes individuals holding posts as consultants to firms. Any individual whose advice is specifically sought by a Health Board in relation to any commercial transaction where such an interest arises must declare that interest. Health Boards should establish and maintain registers both of the financial interests of any staff involved in purchasing/commercial policy and of any 'gifts or considerations' received by staff from any commercial sources."

I would be grateful if you could provide a copy of your last available register for the previous 2 years.

**Appendix 7 - CCG Template Register of Interests**

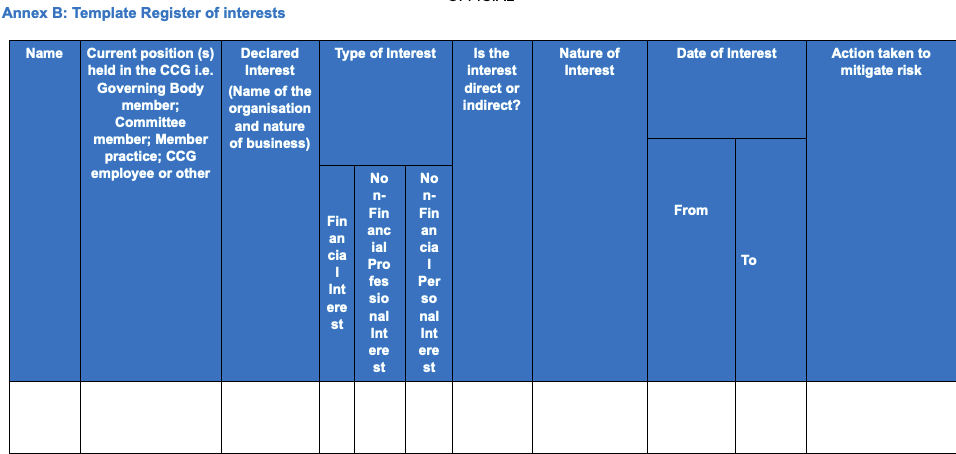

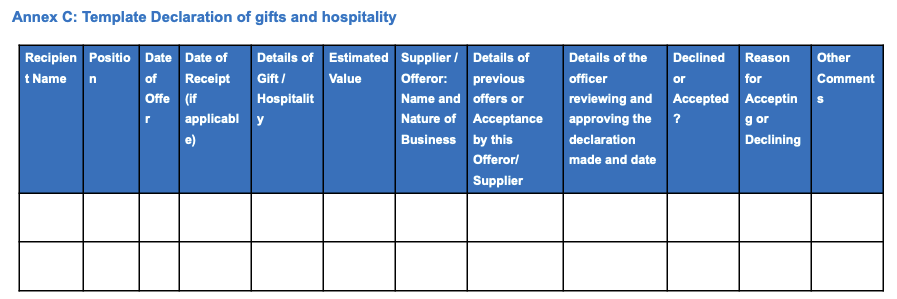

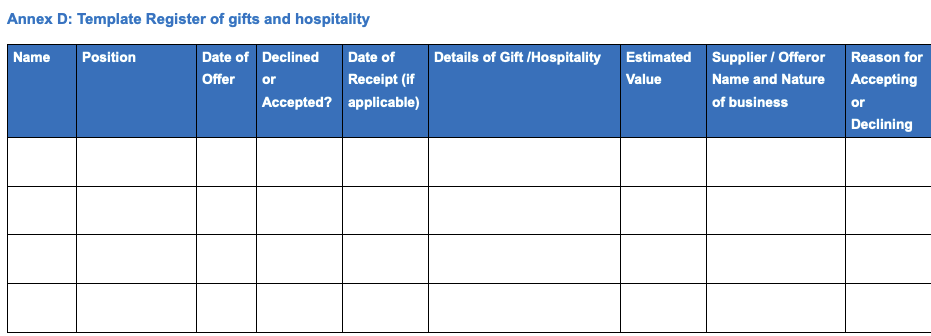

**Appendix 8- Notes on search strategy for CCGs**

1) NHS England has published a standard template for CCGs for staff declaration of interest <https://www.england.nhs.uk/publication/conflicts-of-interest-management-templates/>

These include:

Name, Current Position, Declared Interest, Type of Interest (Financial, Non financial (professional) non financial (personal), whether the interest was direct or indirect, the nature of the interest, the date of the interest, and action taken to mitigate risk. All of these had to be included for the register to be labelled as compliant. Some templates used contained all the requirements of NHSE templates plus additional information.

They have also produced a template for Gifts and Hospitality.

This includes Name, Position, Date of Offer, Declined or Accepted?, Date of Receipt, Details of Gift/Hospitality, Estimated Value, Supplier/Offerer Name and Nature or Business, Action to mitigate against conflict, Reason for Accepting or Declining.

2) In category (5), CCGs had to categorise for financial interest, non financial interest (personal) and non financial interest (professional). If all three were not present (as in the template) they were deemed non compliant.
3) For action to mitigate risk, category (9), this also included a ‘comments’ box as per template C, in Gifts and Hospitality.
4) In category (10), ability to search relates to either a search box, or an alphabetical list. Alphabetisation by first name was not counted as searchable.
5) Category (11), we assessed whether it was possible by design to make a declaration of having no interests; ie by a specific template layout designed to answer ‘no’, or by virtue of people having made a positive declaration that they have no interests to declare.
6) Category (12), Inclusion of a column with request to disclose either pay bands or amounts were included.
7) Category (13), some organisations have amalgamated the gifts and hospitality register and others have not: we recorded whether or not this was the case.
8) Category (14), any listing of a professional declining to disclose e.g. ‘permission not given for publication’, any entry where a sentence relating to disclosure was incomplete or information clearly missing. It was not possible to search each register line by line for retractions and we expect to have missed some.

These were marked by two researchers independently and disagreements resolved by discussion.

We also captured the date of publication and the type of register (e.g Staff Gifts and Hospitality, Governors.) It was in date if published in the last 18 months.

Contacting CCGs: where neither researcher was able to find CCG conflicts of interest and a gift and hospitality register, we contacted the media department of the CCG by email, using the same wording as for NHS Trusts. We requested any publicly available register of interests. If there was no reply in 7 days we wrote again. If there was no response we recorded this. If there was a response we recorded that we required to contact the CCG for registers and then extracted the information as before.
