## Supplementary material for "How are declarations of interest working? A cross sectional study in declarations of interest in medical practice in Scotland and England in 2020/2021": Tables 1-7

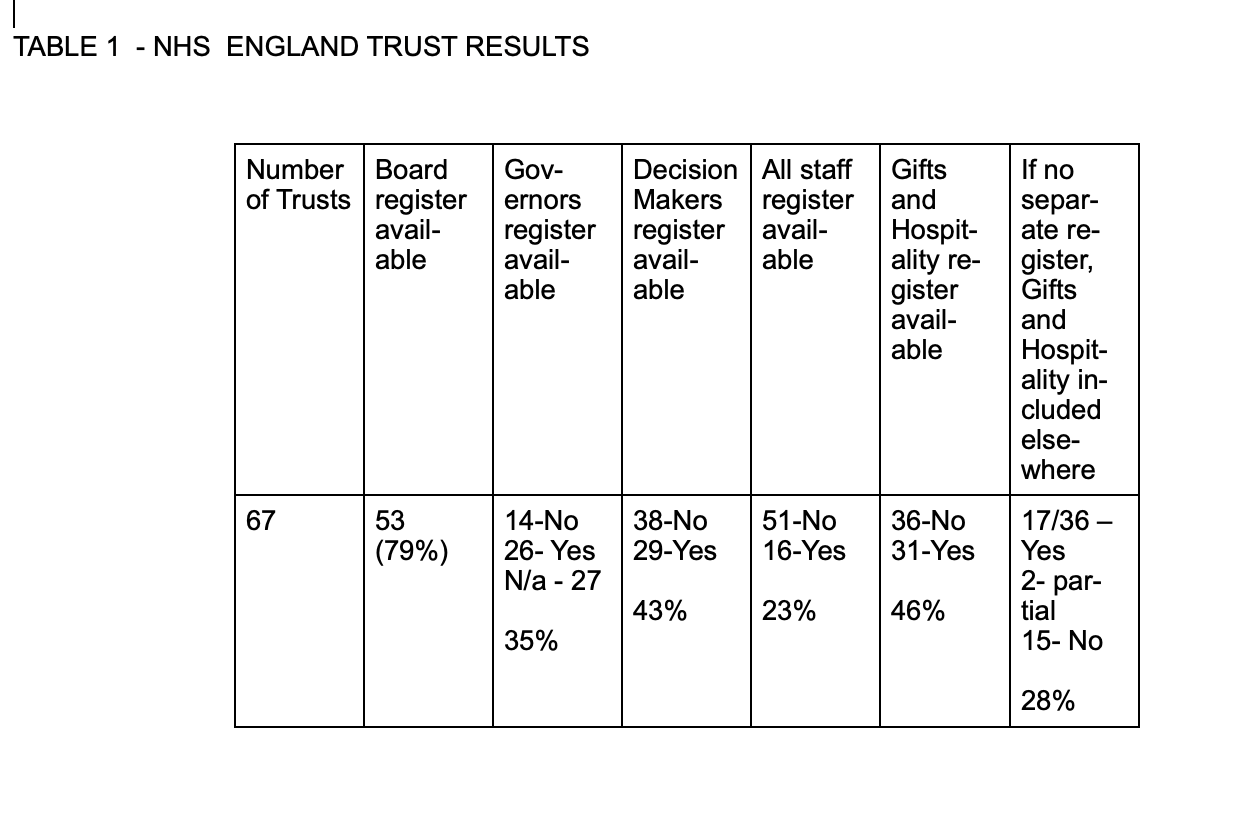

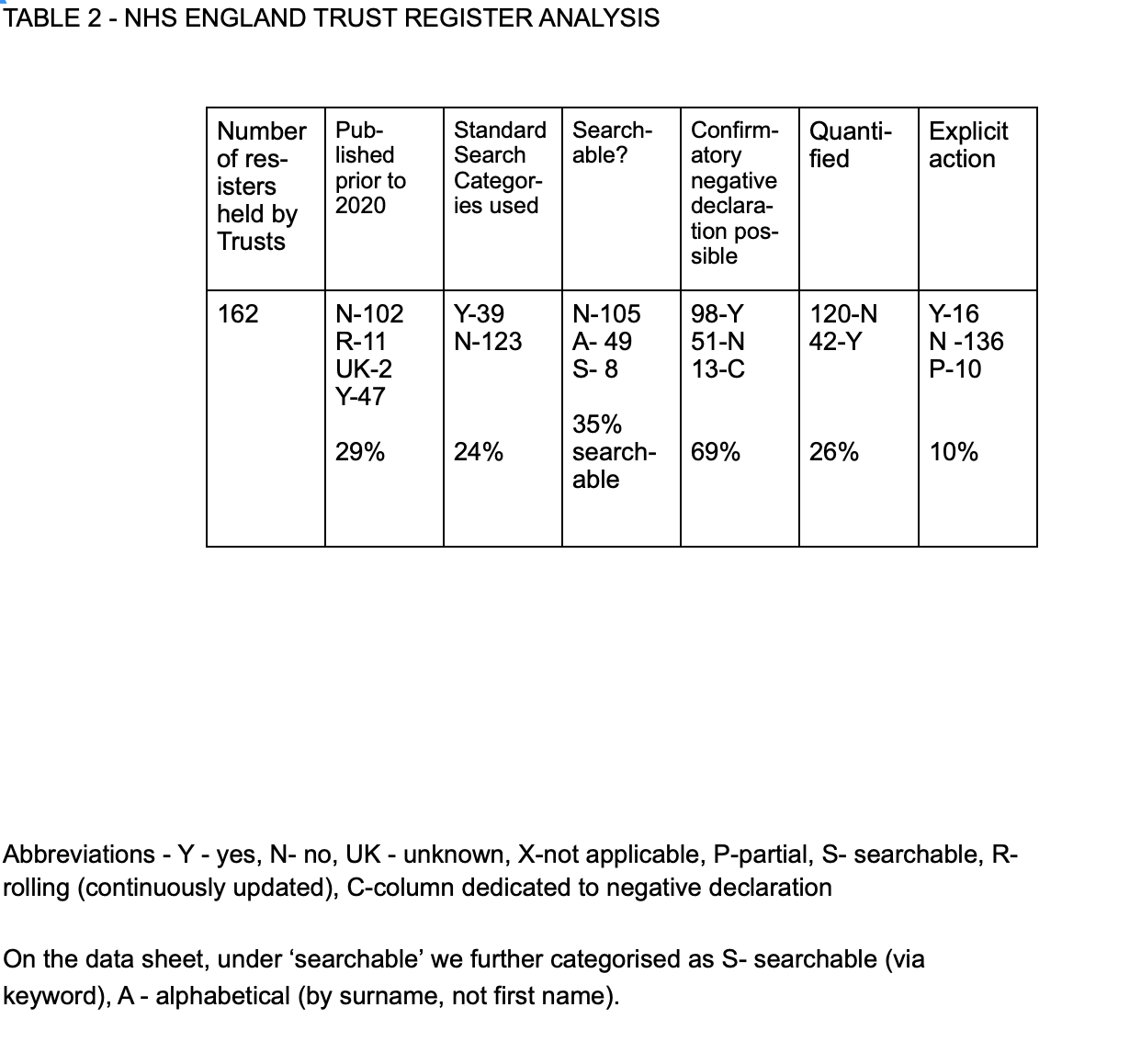

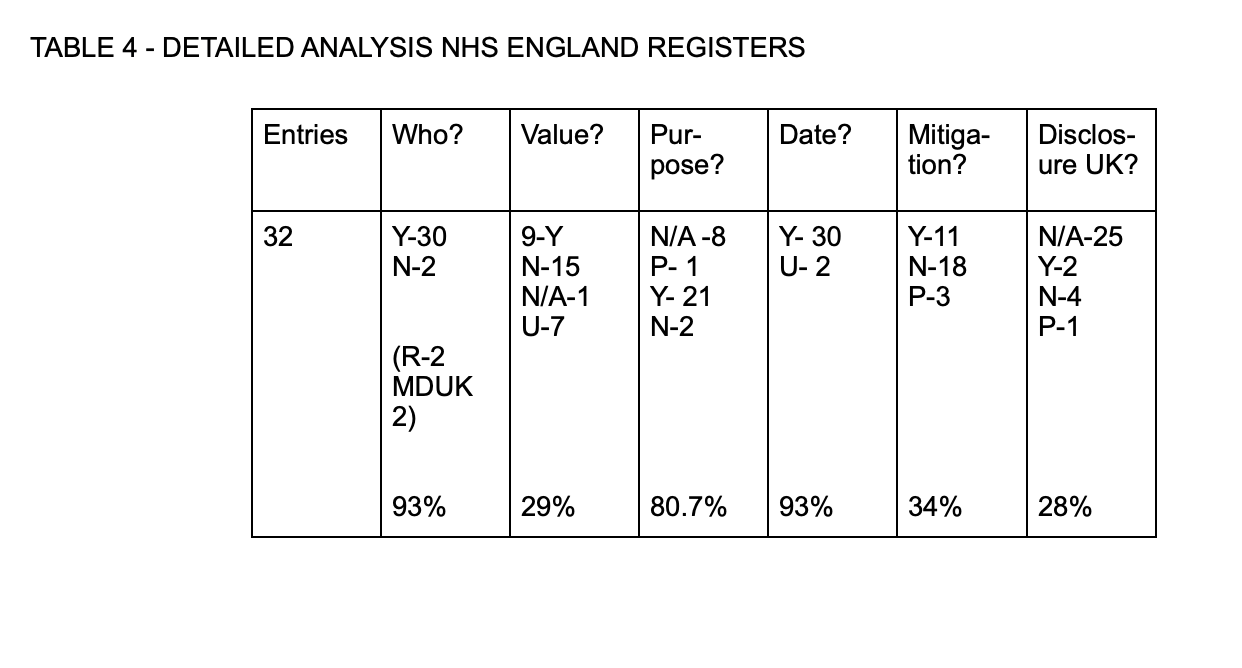

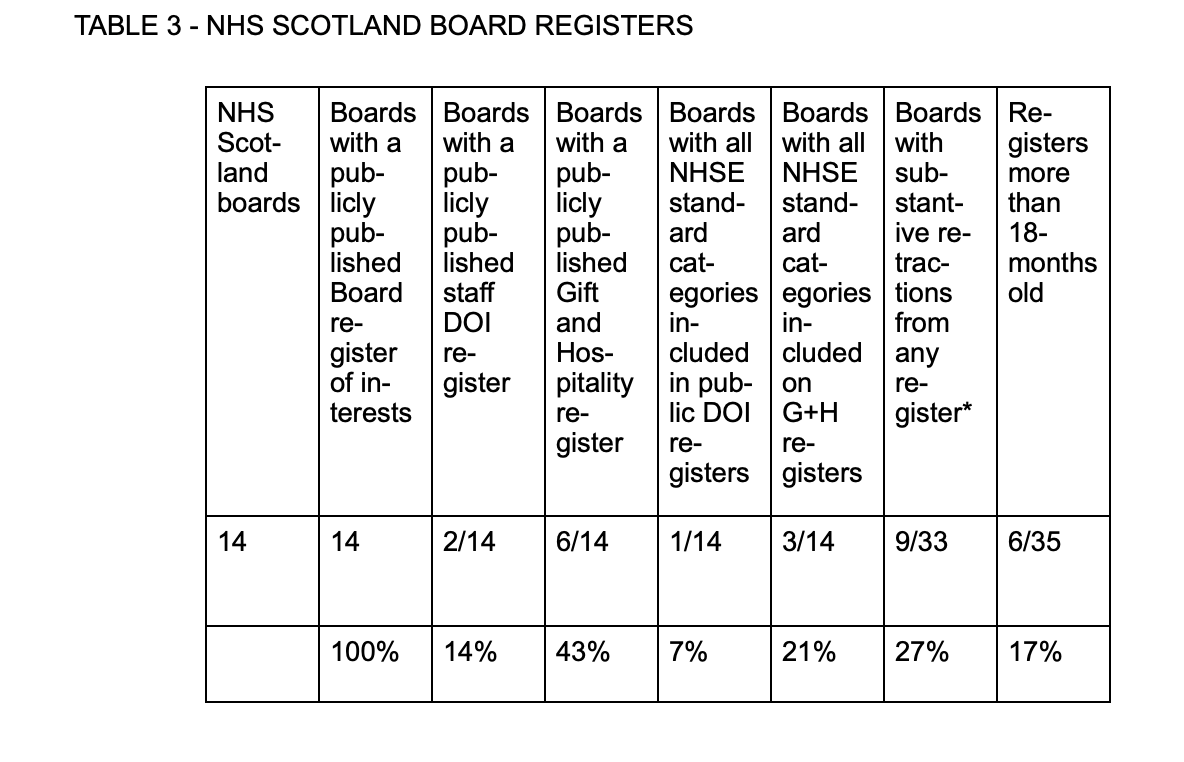

•Footnote to table 3 *Staff home address removal was not included as a redaction
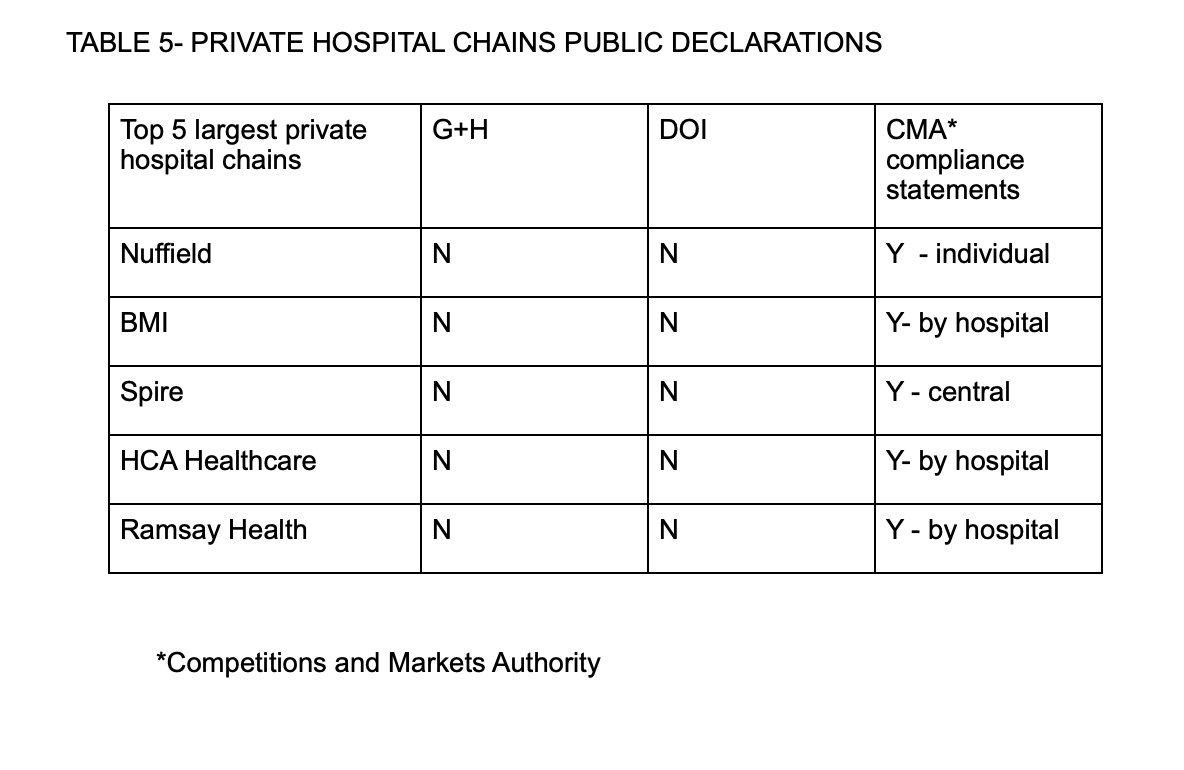

*referrals needed for psychiatric appointments

**referrals done internally from private GP to private consultant

GH - Gifts and Hospitality

DOI - Declaration of Interest

G+H/DOI - Gifts and Hospitality/Declaration of Interest

CMA - Competition and Marketing Authority Statement
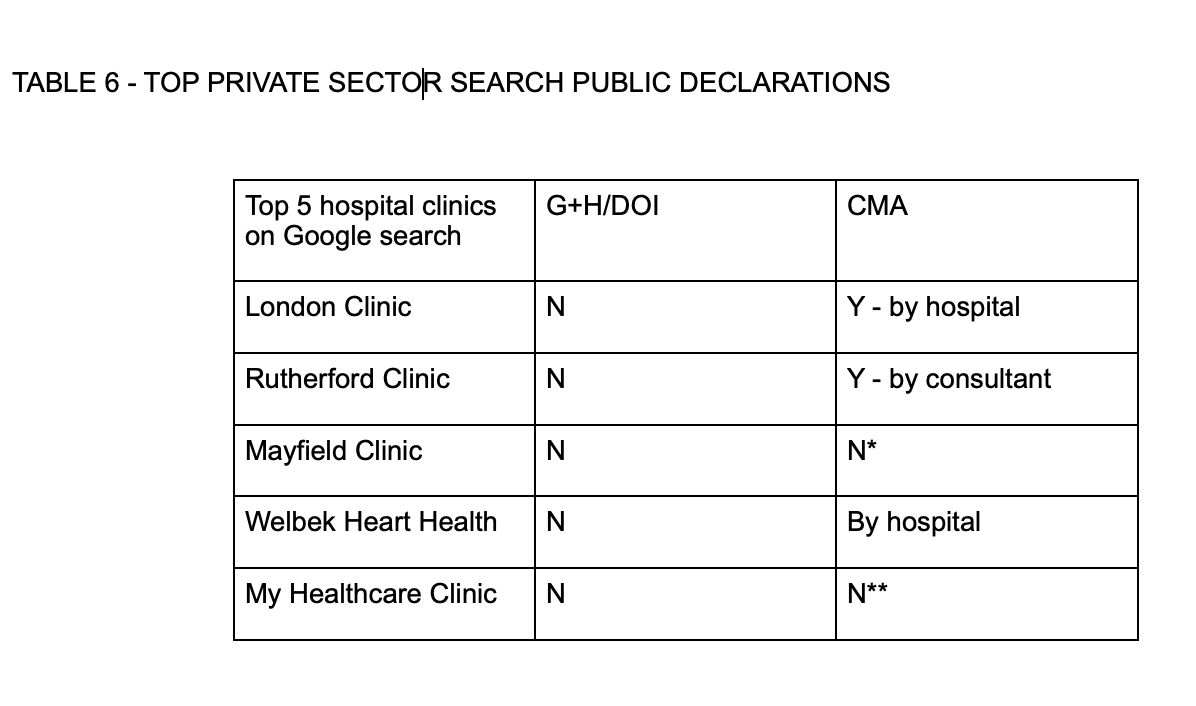

TABLE 7 - CCG REGISTER RESULTS

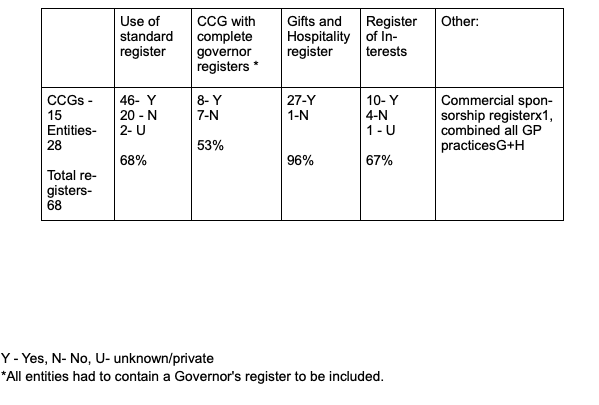
