## Supplementary material for "How are declarations of interest working? A cross sectional study in declarations of interest in medical practice in Scotland and England in 2020/2021": Box 1 and 2

Box 1 - Examples of high quality declarations

“Loyalty Interest - sits on one of the technology appraisal committees at NICE (decides on whether high cost drugs are cost effective for use within the NHS) (unremunerated)."

" Loyalty interest - excluded from the decision making process for the vacant post to avoid conflict of interest, and is two tiers of management above (family member) position." “

“Provision of advice to Janssen about the likely clinical benefits and/or risks of one of their licensed products, Esketamine. The product is licensed for use in the UK, though neither Acute Mental Health Services nor the Trust currently prescribe it (July & Sep 2020). No financial interests in whether this medication is used or not. “

Box 2 - Examples of low quality declarations

"I have been sponsored by pharmaceutical companies to attend Fertility meetings in the past.”

Declaring £1 worth of hospitality: "I have no sight of the costs incurred by hospitality provider and am not prepared to speculate, and have therefore entered an arbitrary low sum. Hospitality was related to comfortable attendance at the meeting only."

“Nine patents currently held, details available upon request.”

“Speaking at various hospitals in China”

“Grant sponsored by industry”
